# Smoking modulates different secretory subpopulations expressing SARS-CoV-2 entry genes in the nasal and bronchial airways

**DOI:** 10.1101/2021.03.30.21254564

**Authors:** Ke Xu, Xingyi Shi, Chris Husted, Rui Hong, Yichen Wang, Boting Ning, Travis Sullivan, Kimberly M Rieger-Christ, Fenghai Duan, Helga Marques, Adam C. Gower, Xiaohui Xiao, Hanqiao Liu, Gang Liu, Grant Duclos, Avrum Spira, Sarah A. Mazzilli, Ehab Billatos, Marc E. Lenburg, Joshua D. Campbell, Jennifer Beane

## Abstract

Coronavirus Disease 2019 (COVID-19) is caused by severe acute respiratory syndrome coronavirus 2 SARS-CoV-2), which infects host cells with help from the Viral Entry (VE) proteins ACE2, TMPRSS2, and CTSL^1–4^. Proposed risk factors for viral infection, as well as the rate of disease progression, include age^5,6^, sex^7^, chronic obstructive pulmonary disease^7,8^, cancer^9^, and cigarette smoking^10–13^. To investigate whether the proposed risk factors increase viral infection by modulation of the VE genes, we examined gene expression profiles of 796 nasal and 1,673 bronchial samples across four lung cancer screening cohorts containing individuals without COVID-19. Smoking was the only clinical factor reproducibly associated with the expression of any VE gene across cohorts. ACE2 expression was significantly up-regulated with smoking in the bronchus but significantly down-regulated with smoking in the nose. Furthermore, expression of individual VE genes were not correlated between paired nasal and bronchial samples from the same patients. Single-cell RNA-seq of nasal brushings revealed that an ACE2 gene module was detected in a variety of nasal secretory cells with the highest expression in the C15orf48+ secretory cells, while a TMPRSS2 gene module was most highly expressed in nasal keratinizing epithelial cells. In contrast, single-cell RNA-seq of bronchial brushings revealed that ACE2 and

TMPRSS2 gene modules were most enriched in MUC5AC+ bronchial goblet cells. The CTSL gene module was highly expressed in immune populations of both nasal and bronchial brushings. Deconvolution of bulk RNA-seq showed that the proportion of MUC5AC+ goblet cells was increased in current smokers in both the nose and bronchus but proportions of nasal keratinizing epithelial cells, C15orf48+ secretory cells, and immune cells were not associated with smoking status. The complex association between VE gene expression and smoking in the nasal and bronchial epithelium revealed by our results may partially explain conflicting reports on the association between smoking and SARS-CoV-2 infection.

## Main

As of February 14^th^, 2021, over 108 million confirmed cases and 2.39 million deaths have been reported globally for COVID-19 (https://www.who.int/emergencies/diseases/novel-coronavirus-2019/situation-reports). SARS-CoV-2 infects cells by utilizing host cell-surface proteins for viral entry (VE). VE proteins include angiotensin-converting enzyme 2 (ACE2), which provides a binding site for the virus; proteases such as transmembrane protease serine 2 (TMPRSS2) that can cleave the viral S glycoprotein; and cathepsin L (CTSL), which can also aid with S glycoprotein priming^1–4^. Prior studies have attempted to establish associations between the expression of VE genes and clinical variables to identify factors that may influence infection incidence or COVID-19 severity.

However, some conflicting findings have been reported. Smith et al^14^, Wang et al^15^, and Brake et al^16^ reported that the expression level of ACE2 in the tracheal epithelium is elevated with cigarette smoke, while Zhang et al^18^ and Aliee et al^19^ reported that the expression level of ACE2 is elevated with smoking only in the small airway epithelium but not in the large airway epithelium. Bunyavanich et al^17^ reported that ACE2 expression levels were associated with age but did not control for other important clinical variables; however, Smith et al^14^ found that ACE2 expression was equivalent between young and elderly individuals. Moreover, earlier studies with limited numbers of subjects led to the conclusion that race was associated with ACE2 expression^20^, which was not found by Smith et al in a larger dataset^14^. Lastly, lung airway expression of both ACE2 and TMPRSS2 was found to be significantly up-regulated in patients with chronic obstructive pulmonary disease (COPD) compared with healthy subjects or in smokers compared with non-smokers^21^, but these associations were found without adjusting for other important factors such as age and sex. The goal of this study is to resolve these discrepancies by studying gene expression profiles in a large number of airway samples from multiple cohorts and to compare and contrast the biological processes associated with the expression of VE genes in the nasal and bronchial compartments.

Gene expression profiles of 796 nasal and 1,673 bronchial brushing samples were generated using bulk RNA-seq from 4 cohorts, DECAMP^22^ (Detection of Early Lung Cancer Among Military Personnel), AEGIS^23^ (Airway Epithelial Gene expression in the Diagnosis of Lung Cancer), BCLHS^24^ (British Columbia Lung Health Study and Pan-Canadian Lung Health Study), and PCA^25^ (Pre-Cancer Atlas) (**Table 1**). Study participants were current or former smokers undergoing bronchoscopy as part of either a diagnostic workup or a screening process for lung cancer. Clinical data collected on samples included smoking status, sex, age, forced expiratory volume during the first second (FEV1) percent Predicted, and, in the AEGIS study, lung cancer status (**Supplemental Tables 1**). Notably, we observed different patterns of association with smoking between ACE2 and TMPRSS2 gene expression between the nasal and bronchial epithelium. ACE2 was down-regulated in the nasal epithelium of current smokers in 1 out of 2 cohorts (p < 0.01) but strongly up-regulated in the bronchus in all four cohorts (p < 0.01; **Figs. 1a and 1b; Supplemental Fig. 1a and 1b**). TMPRSS2 expression was not associated with smoking in the nose but its expression was up-regulated in current smokers in the bronchus in 3 out of 4 cohorts (p < 0.001). CTSL expression was down-regulated in current smokers in the nasal epithelium in 1 out of 2 cohorts (p < 0.01) and the bronchial epithelium in 3 out of 4 cohorts (p < 0.05). These genes were not strongly associated with other clinical variables (sex, age, FEV1% Predicted) across cohorts in either the nasal or bronchial samples in contrast to some previous studies^14,17,20,21^. Adjusting for lung cancer diagnosis in the AEGIS cohort did not change these observed correlations, although positive lung cancer status negatively correlated with ACE2 expression in the nasal epithelium (p<0.05, **Supplemental Fig. 1b**). A lack of correlation in the expression of each VE gene between the nose and bronchus was observed in paired samples from the same subjects (p > 0.05; **Fig. 1c**; **Supplemental Fig. 1c, Supplemental Table 3**), suggesting that the biological processes that influence the expression of these genes may differ across airway sites.

**Table 1.** Summary of Patient Demographics. Definition of abbreviations: DECAMP = Detection of Early Lung Cancer Among Military Personnel, AEGIS = Airway Epithelial Gene expression in the Diagnosis of Lung Cancer, BCLHS = British Columbia Lung Health Study and Pan-Canadian Lung Health Study, PCA = Pre-Cancer Atlas. Values are presented as mean (SD).

|  | DECAMP<br>Nose | AEGIS<br>Nose | DECAMP<br>Bronchus | AEGIS<br>Bronchus | BCLHS<br>Bronchus | PCA<br>Bronchus |
| --- | --- | --- | --- | --- | --- | --- |
| **Total number of samples/cells** | 288 | 505 | 360 | 938 | 238 | 137 |
| **Smoking Status (%)** |  |  |  |  |  |  |
| Current | 125 (43.40) | 186 (36.83) | 153 (42.5) | 442 (47.12) | 99 (41.6) | 77 (56.2) |
| Former | 156 (54.17) | 319 (63.17) | 202 (56.11) | 496 (52.88) | 139 (58.4) | 60 (43.8) |
| Never | 0 | 0 | 0 | 0 | 0 | 0 |
| Missing | 7 (2.43) | 0 | 5 (1.39) | 0 | 0 | 0 |
| **Sex, n (%)** |  |  |  |  |  |  |
| Male | 219 (76.04) | 317 (62.78) | 282 (78.33) | 584 (62.26) | 135 (56.72) | 70 (51.09) |
| Female | 69 (23.96) | 188 (37.23) | 78 (21.67) | 354 (27.74) | 103 (43.28) | 67 (48.91) |
| Missing | 0 | 0 | 0 | 0 | 0 | 0 |
| **Age** |  |  |  |  |  |  |
| Mean (SD) | 67 (8) | 60 (11) | 65 (7) | 63 (11) | 65 (6) | 58 (7) |
| Missing | 0 | 0 | 0 | 0 | 0 | 0 |
| **FEV1 (% predicted)** |  |  |  |  |  |  |
| Mean (SD) | 75.32 (19.15) | 73.00 (21.28) | 72.71 (20.26) | 68.73 (22.18) | 80.86 (20.68) | 74.32 (20.72) |
| Missing | 28 | 355 | 0 | 633 | 0 | 4 |
| **Having Lung Cancer (%)** |  |  |  |  |  |  |
| Yes | NA | 309 (61.19) | NA | 710 (75.69) | NA | NA |
| No | NA | 196 (38.81) | NA | 228 (24.31) | NA | NA |
| Missing | NA | 0 | NA | 0 | NA | NA |

**Figure 1.**
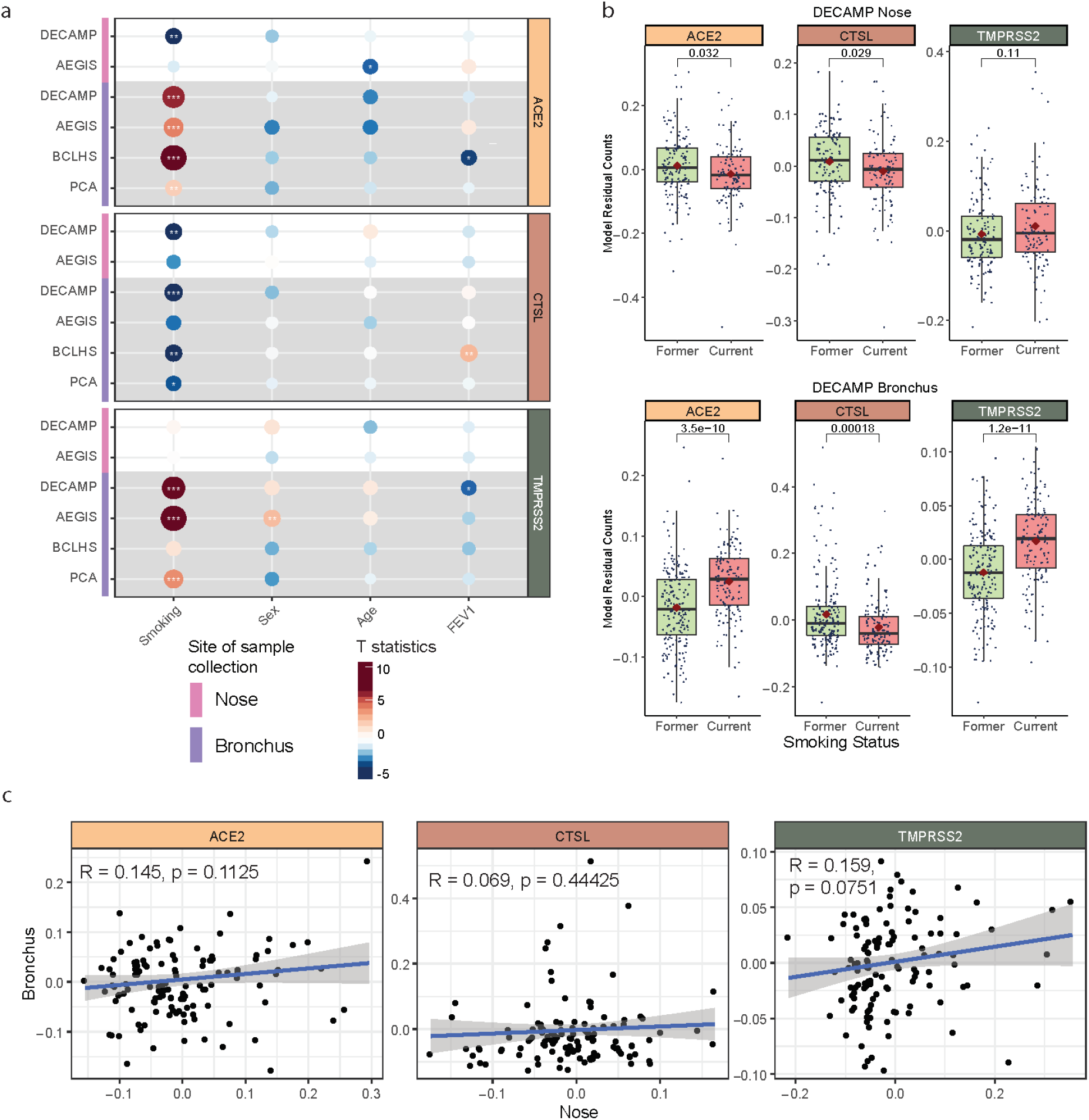
ACE2, CTSL, and TMPRSS2 gene expression is strongly associated with current cigarette smoking in both the nasal and bronchial epithelium. (**a**) Associations between the expression of ACE2 (tan), CTSL (brown), and TMPRSS2 (olive) and clinical covariates in the nasal and bronchial epithelium. Nasal (pink) and bronchial (purple) brushings were collected from subjects at high risk of developing lung cancer from four different studies (DECAMP bronchus: N = 341, AEGIS bronchus: N = 305, BCLHS bronchus: N = 238, PCA bronchus: N = 133, DECAMP nose: N = 253, AEGIS nose: N = 150). The size and color of the bubbles represent the significance and magnitude, respectively, of the *t* statistic calculated using linear modeling of VE gene expression as a function containing the four clinical variables, correcting for batch and mean Transcript Integrity Number (mTIN). Significance levels: * *p* < 0.05, ** *p* < 0.01, *** *p* < 0.001. (**b**) Boxplots showing the significance of associations between VE expression and smoking status in the DECAMP cohort, assessed by Student’s *t* test on residual counts after correction for sex, age, percentage of predicted FEV1, batch, and mTIN (*p* < 0.05). (**c**) Expression of VE genes is not significantly correlated (*p* > 0.05, Pearson correlation) between paired DECAMP nasal (x-axis) and bronchial (y-axis) epithelial samples (N = 123). Residual counts adjusted for sex, age, percentage of predicted FEV1, batch, and mTIN were used for the comparison. The blue line is the line of best fit and the gray shading represents the 95% confidence level interval for predictions from the linear model.

To further explore the biological pathways associated with VE genes in each site of the airway, we developed co-expression networks using Weighted Gene Correlation Network Analysis (WGCNA)^26^. We identified 58 and 48 gene modules within the nasal and bronchial cohorts, respectively (**Fig. 2a** and **2b, Supplemental Fig. 2; Supplemental Table 2**). The consensus modules eigengenes containing the VE genes significantly correlated with their respective VE gene expression within each cohort (**Supplemental Fig. 3a**, Pearson correlation, p < 2.2E-16, average R = 0.584), suggesting that the eigengene can be used as a surrogate for each individual VE gene. The VE gene module eigengenes showed similar associations with the clinical covariates (**Supplemental Fig. 3b**). While the nasal VE gene module eigengenes were not highly correlated, ACE2- and TMPRSS2-associated module eigengenes in the bronchus were highly correlated across all four datasets (**Supplemental Figs. 2a** and **2b**), suggesting that similar biological processes control these two gene modules in the bronchial compartment.

**Figure 2.**
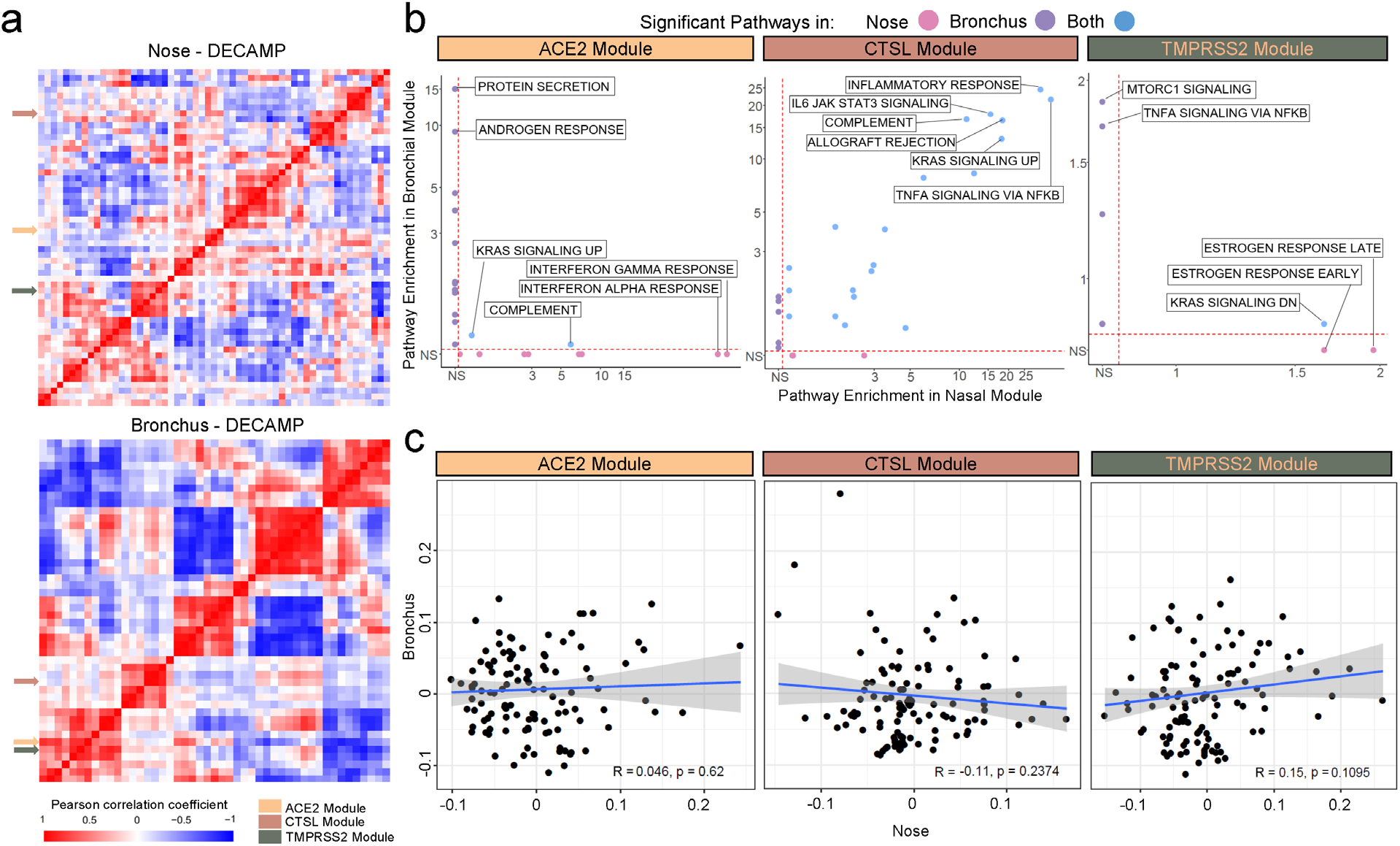
Consensus nasal and bronchial gene co-expression modules containing the VE genes are associated with different biological pathways and are not correlated between tissues. (**a**) WGCNA was used to identify consensus co-expression modules in nasal and bronchial samples from the DECAMP cohort (n=58 and 48 modules, respectively). A heatmap of the correlation of module eigengenes computed from each module demonstrates that ACE2 and TMPRSS2 modules were more highly correlated with each other in the bronchus than in the nose. The CTSL module was not correlated with ACE2 or TMPRSS2 modules in either the nose or bronchus. (**b**) Scatterplots comparing the overrepresentation of MSigDB Hallmark pathway gene sets in each VE module in the nose and bronchus. The −log_10_(FDR *q*) values denoting the degree of overrepresentation of each gene set in each module in the nose and bronchus are shown on the x- and y-axes, respectively. The overrepresentation of gene sets in the ACE2 and TMPRSS2 modules is largely discordant between the nose and bronchus, whereas several immune-related pathways are overrepresented in the CTSL module in both sites. (**c)** VE module eigengenes were not significantly correlated (Pearson *p* > 0.05) between paired nasal (x-axis) and bronchial (y-axis) samples in DECAMP (N = 114). The blue line is the line of best fit and the gray shading represents the 95% confidence level interval for predictions from the linear model.

Comparing biological pathways enriched among VE gene modules between the nose and bronchus revealed (**Fig. 2c**; **Supplemental Table 3**) that genes of the nasal ACE2 module were enriched in the inflammatory response, interferon-alpha signaling, and interferon-gamma signaling pathways while the bronchial ACE2 gene module was enriched in genes associated with protein secretion and androgen response. Genes of the TMPRSS2 module in the nose were enriched in estrogen response and KRAS signaling pathways while the module in the bronchial epithelium was enriched in MTORC1 and TNFα-NFκB signaling pathways. The lack of shared biological pathways mirrors the small number of genes shared by the nasal and bronchial modules for ACE2 (n = 5) and TMPRSS2 (n = 12). In contrast, the nasal and bronchial CTSL modules shared 177 genes (Fisher’s exact test, p = 2.23E-10) and were both enriched in genes related inflammatory response and TNF-alpha signaling pathways. Furthermore, in the bronchus, ACE2 and TMPRSS2-associated modules share enrichment in genes involved in the p53 pathway, adipogenesis, androgen response, MTORC1 signaling, and estrogen response (**Supplemental Fig. 3c**). We also did not observe significant correlations between the eigengenes of each VE gene module in the paired nasal and bronchial samples (**Fig. 2d** and **Supplemental Fig. 3d**). The VE gene module and single-gene analyses are consistent and suggest that there are important differences between VE gene expression and biology between the upper and lower airways.

In order to determine if different cell populations contribute to the difference in VE gene expression between airway compartments, we leveraged single-cell RNA sequencing (scRNA-seq) data to characterize the expression patterns of individual VE genes and VE gene modules across major epithelial and immune cell types. We profiled 34,833 cells from 9 nasal brushings (4 collected from two volunteers and 5 from 5 patients undergoing lung cancer screening) and 2,075 cells from 17 bronchial brushings from patients undergoing bronchoscopy for suspicion of lung cancer (**Supplemental Tables 6, 7**). A total of 17 and 15 transcriptionally distinct cell clusters were identified in the nose and bronchus, respectively, many of which expressed similar marker genes: KRT5, KRT15 (basal cells); FOXJ1, C20orf85, CDC20B (ciliated cells); SCGB1A1, SERPINB3 (club cells); MUC5AC, TFF1, TFF3 (goblet cells); FOXI1, CFTR (ionocytes); CD3D, CD8A (T cells). We also identified two bronchial goblet-like secretory populations: one characterized by high expression of CEACAM5 but not MUC5AC, previously described as peri-goblet cells^27^, and the other by high expression of HLA-DQA1 and MHC class II genes (**Figs. 3a-b, Supplemental Fig. 4**). In the nose, we discovered two club-cell-like secretory cell clusters (STATH+ and C15orf48+), as well as a novel cell cluster of “keratinizing epithelial cells” expressing genes involved in cornification, epidermis development, and keratinocyte differentiation, such as SPRR3 and SPRR2A. Immune populations represent <1% and 26% of total cell populations in the nasal and bronchial epithelium, respectively.

**Figure 3.**
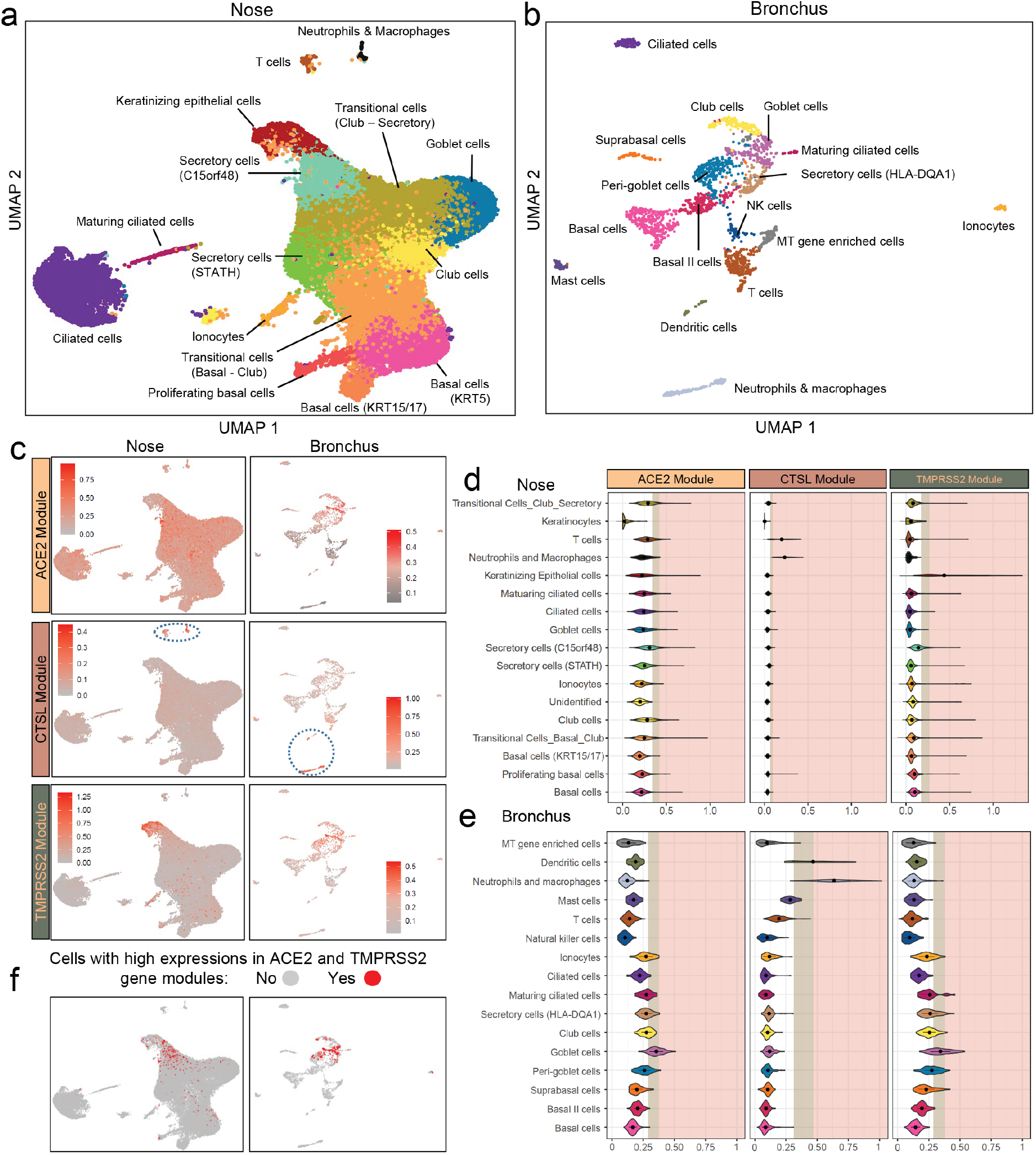
Expressions of VE gene modules among various cell types. Single-cell RNA-seq was performed (**a**) on nasal brushings from 7 patients (n=34,833 cells) and (**b**) bronchial brushings from 17 patients (n=2,075 cells). Cell types were inferred from the expression of known marker genes. **(c)** UMAP projections showing the expression pattern of VE gene modules (ACE2, top; CTSL, middle; TMPRSS2 bottom) across different cell types in the nasal (left) and bronchial (right) epithelium. The cells are colored gray for low expression and red for high expression of metagene scores of each VE gene module. Immune cells with high expression of the CTSL modules were encircled in blue (**d-e**). Violin plot showing the metagene score for each VE gene module across the cell types in (**d**) nasal and (**e**) bronchial epithelium. For each violin plot, metagene expression is designated as elevated (light brown) or highly elevated (pink) if it is greater than one or two standard deviation above the mean metagene score, respectively. (**f**) UMAP projections showing cells greater than 1 standard deviation above the mean for both ACE2 and TMPRSS2 modules (i.e., double positive). Double positive cells were overrepresented in nasal secretory (C15orf48+) and keratinizing epithelial cells (left) and bronchial goblet cells (right).

We found low expression of VE gene in the single cell data, consistent with other reports (**Supplemental Fig. 5**)^28,29^, so we calculated VE gene module metagene scores in each cell to understand how biological processes associated with the VE genes in the bulk RNA-seq data are distributed across nasal and bronchial cell populations (**Figs. 3c-e**). In the nose, the ACE2 module was moderately expressed across many cell types but was most highly expressed in C15orf48+ secretory cells and club cells. The TMPRSS2 module was predominantly expressed in the keratinizing epithelial cells, followed by C15orf48+ secretory cells. The expression of gene modules of ACE2 and TMPRSS2 is different from the gene expression pattern reported by Sungnak et al in that TMPRSS2 gene expression was limited to ACE2+ cells^30^. In the bronchus, both the ACE2 and TMPRSS2 modules were expressed at the highest levels in the goblet cell population, in agreement with previous findings^31^. The highest median metagene score for the CTSL module was in the neutrophils and macrophages in both the nose and bronchus; however, expression was also high in T cells in the nose and dendritic cells in the bronchus. We found very few cells co-expressing ACE2, CTSL, and TMPRSS2 at high levels – 114 out of 34833 cells in the nose and no cells in the bronchus. On the other hand, we found that ACE2 and TMPRSS2 modules were more like to be highly co-expressed in nasal keratinizing epithelial cells (odds ratio = 7.56), nasal C15orf48+ secretory cells (N = 518, odds ratio = 7.36), and bronchial goblet cells (odds ratio = 16.80) (N = 806, **Supplemental Table 6**, FDR q < 0.001). Thus, ACE2 and TMPRSS2 were found to be expressed in different nasal and bronchial cell populations, which may be influenced by smoking in ways that lead to their divergent correlation with smoking in the two airway compartments.

To investigate how smoking modulates different cell populations, we computationally deconvolved cell population proportions in the bulk RNA-seq data using gene markers identified from the single-cell data (**Supplemental Table 7**). Higher proportions of goblet cells were observed in current smokers in both the nose and bronchus in all 6 cohorts, consistent with previous studies^27^. Increased proportions of ionocytes in current smokers were also observed in the nose (1 out of 2 cohorts) and bronchus (all 4 cohorts). The proportion of ciliated cells was significantly lower in smokers in all 6 cohorts (**Figs. 4a-b, Supplemental Figs 6a-d**, p < 0.05). While goblet cell proportions were significantly higher in smokers in both the nose and bronchus, the ACE2 module was highly expressed only in bronchial, not nasal goblet cells. These results may explain the lack of association of ACE2 expression with smoking in the bulk RNA-seq data. Additionally, in the AEGIS nasal data (**Supplemental Fig. 6**) the fraction of keratinizing epithelial cells is increased and that of STATH+ secretory cells is decreased in current smokers, suggesting that shifts in these populations may be partially responsible for the differential correlation with smoking between ACE2 and TMPRSS2 in the nose. With respect to CTSL, both nasal brushings showed a significant decrease in the proportion of the neutrophil/macrophage population with respect to smoking. While this may partially explain the down-regulation of CTSL in smokers in the nasal bulk RNA-seq data, the overall proportions of these populations estimated in the bronchial data by the deconvolution were close to zero for most samples, which could have prevented us from confirming the decrease of this population with smoking in the bronchus.

**Figure 4.**
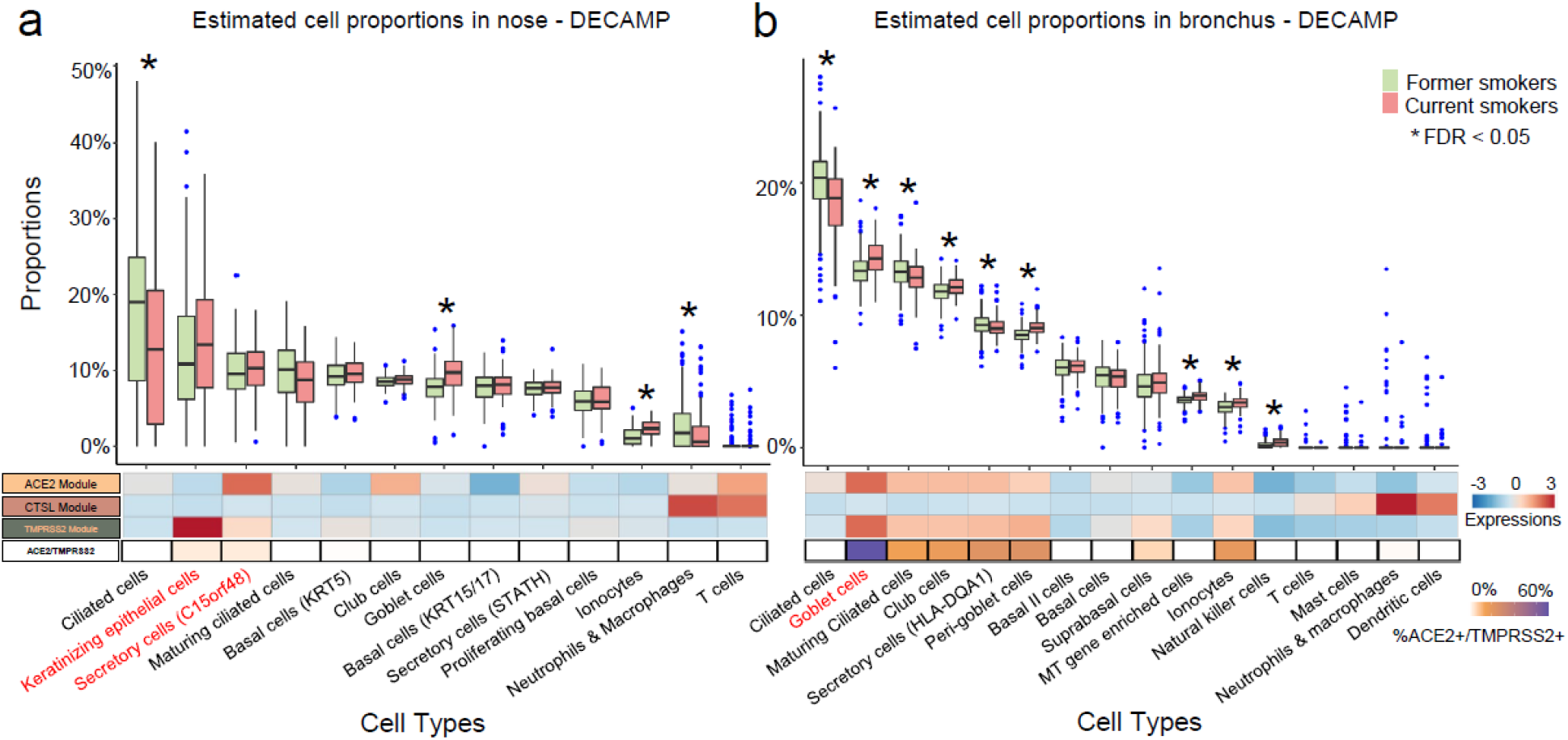
Deconvolution of bulk RNA-seq data shows increased proportions of goblet cells and ionocytes in both nasal and bronchial epithelium of current smokers compared to former smokers. (**a-b**) Top: boxplots of cell type proportions estimated by AutoGeneS in bulk RNA-seq data from nasal (N = 281) and bronchial brushings (N = 355) obtained from current and former smokers in the DECAMP cohort. Significant cell proportion differences between current and former smokers were determined by Wilcoxon test (* indicates FDR q < 0.05). Bottom: the heatmap displays the average VE module score for each cell population from the single-cell RNA-seq data (first three rows), as well as the proportion of each cell type that is ACE2+/TMPRSS2+ (i.e., the expression of both genes is one standard deviation above the average). Cell types that express both ACE2 and TMPRSS2 modules are in red. While goblet cell proportions were significantly higher in smokers in both the nose and bronchus, the ACE2 module was only highly expressed in goblet cells from the bronchus, which may explain the lack of association of ACE2 expression with smoking in the bulk RNA-seq data.

The current study investigated the relationship between the expression of genes encoding proteins important for SARS-CoV-2 entry (ACE2, CTSL, and TMPRSS2) and clinical factors including age, sex, COPD, and smoking in both the nose and bronchus. In our datasets, only smoking status showed a consistent association with the expression of VE genes across cohorts within each airway compartment. In the bronchus, current smoking status was associated with higher ACE2 and TMPRSS2 expression; however, in the nose, current smoking status was inversely correlated with ACE2 expression and was not associated with TMPRSS2 expression as in prior studies ^14-16,19^. The cohorts used in this study contain subjects at high risk for developing lung cancer and are thus composed of older subjects (mean age = 63, SD = 8); therefore, we did not find an association between nasal ACE expression and age as previously reported by Bunyavanich et al^17^ in a study comparing children and adults. We also did not find sex to be associated with ACE2 expression as was reported by another study with a lower sample size^18^ and could not confidently assess differences in expression profiles between racial subgroups as our cohorts are predominantly (> 70%) composed of White subjects. Our analyses did not find significant VE gene expression correlations between paired nasal and bronchial samples suggesting that there are differences in biological pathways or cell types between the airway compartments.

Our WGCNA analysis demonstrated that genes co-expressed with ACE2 and TMPRSS2 in the bulk RNA-seq data were more highly correlated to one another in the bronchus than in the nose and that pathway enrichment was different for these modules in the upper and lower airways. Similarly, scRNA-seq data showed that ACE2 and TMPRSS2 modules were co-expressed in bronchial goblet cells but had distinct patterns of expression across nasal cell populations. Our high-resolution nasal scRNA-seq dataset containing over 30,000 cells allowed us to differentiate between secretory cell populations to show that C15orf48+ and keratinizing epithelial cells had the highest expression of ACE2 and TMPRSS2 modules, respectively. In contrast, we found that bronchial goblet cells had the highest expression of both ACE2 and TMPRSS2 modules and only the goblet cell population was increased in smokers in our deconvolution analysis. Of note, ACE2 and TMPRSS2 modules were also co-expressed highly within the peri-goblet cells in the bronchus, which was also observed by Lukassen et al^29^.

Interestingly, the nasal and bronchial CTSL modules showed enrichment of immune-associated biological programs and were highly expressed in similar immune cell populations. Despite these similarities, CTSL gene or module expression was not correlated between paired bronchial and nasal samples potentially due to the low abundance of immune populations in the scRNA-seq and bulk RNA-seq data.

Our study leveraged bulk and single-cell gene expression profiles from large cohorts to show that current cigarette smoking status is consistently associated, in a site-specific manner, with the expression of genes required for SARS-CoV-2 entry in the nose and bronchus. Future work investigating other putative viral entry genes such as FURIN and ATRNL1^29–31^ may build upon these findings to enhance our understanding of the effect of smoking on SARS-CoV-2 infection. The results of our study of the expression of ACE2 and TMPRSS2 suggest that smoking is unlikely to impact the likelihood of SARS-CoV-2 infection in the upper airways, but that it may play a significant role in COVID-19 disease progression and severity.

## Data Availability

All data are either available online or currently being processed to be published on Gene Expression Omnibus (GEO).

https://www.ncbi.nlm.nih.gov/geo/query/acc.cgi?acc=GSE80796

https://www.ncbi.nlm.nih.gov/geo/query/acc.cgi

https://www.ncbi.nlm.nih.gov/geo/query/acc.cgi

## Acknowledgements

We would like to thank Yuriy Alekseyev, Ashley LeClerc, and Kangning Zhang of the Single Cell Sequencing Core at Boston University School of Medicine for their help with data generation.

## Authors contributions

K. X., X. S., C. H., contributed equally to this work (co-first authors); J. C., and J. B. contributed equally to this work (co-senior authors). K. X. is the guarantor of this paper and takes full responsibility for the integrity of the work as a whole. A. S., M. E. L., E. B., J. C., J. B., S. M. initiated the study design. F. D., H. M., and A. C. G. prepared patient characteristics for the DECAMP cohort; K. X., T. S., K. R-C., X. X., H. L., G. L., and G. D. collected and processed samples; K. X., X. S., C. H., R. H., Y. W., and B. N. analyzed data. K. X., X. S., and C. H. prepared the figures; K. X., X. S., C. H., M. E. L., J. C. and J. B. interpreted results; K. X., X. S., and C. H drafted manuscript; K. X., X. S., C. H., M. E. L., J. C., and J. B. edited and revised manuscript; all the others approved the final version of manuscript.

