## Supplemental figures and method for "Smoking modulates different secretory subpopulations expressing SARS-CoV-2 entry genes in the nasal and bronchial airways"

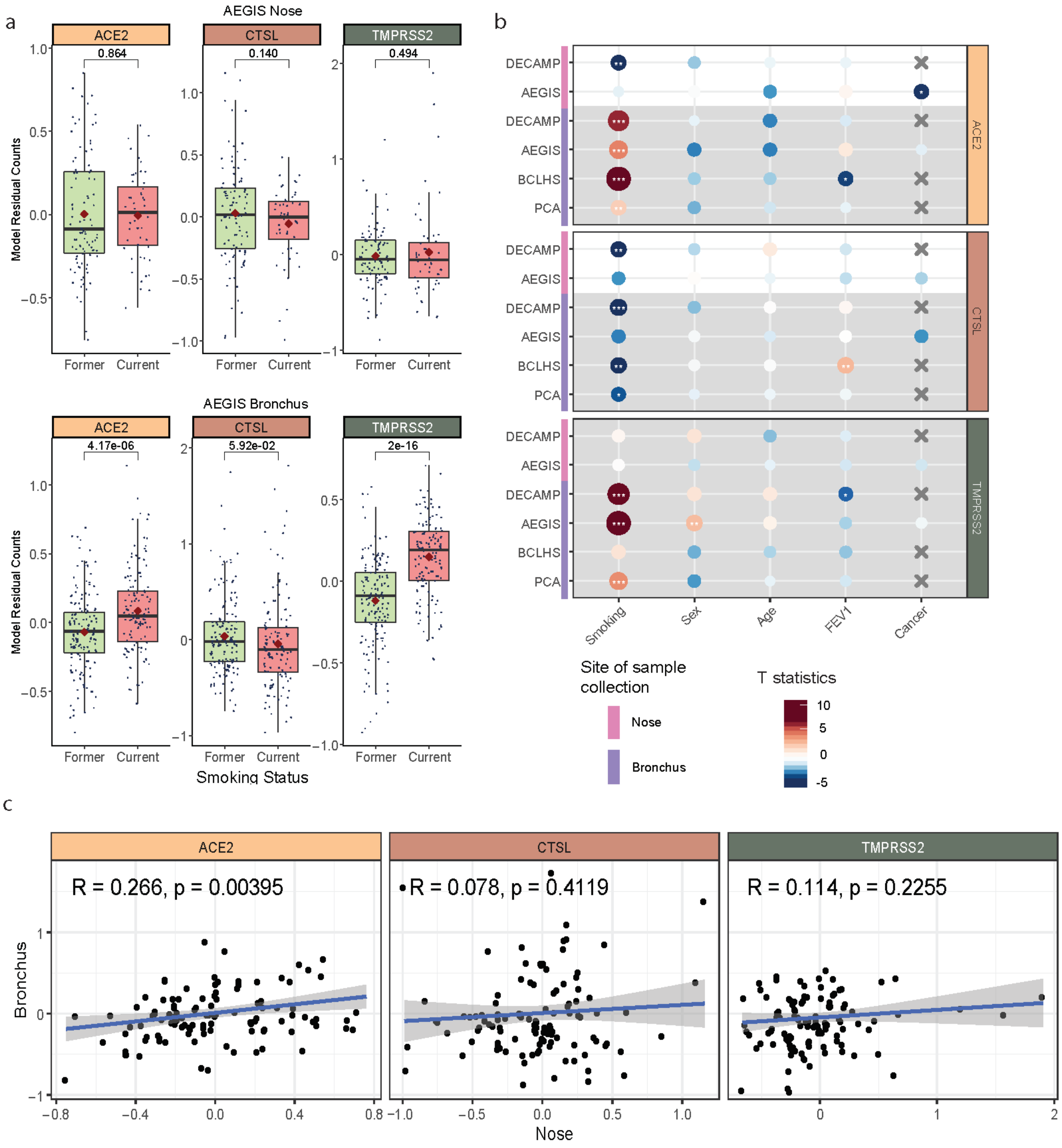


**Figure S1. ACE2, CTSL, and TMPRSS2 expression associated with current cigarette smoking in nasal and bronchial epithelium.**

(**a**) Boxplots showing the significance of associations between VE gene expression and smoking status in nasal (N = 150) and bronchial (N = 305) epithelium from the AEGIS cohort, assessed by Student's t-test on residual expression values after correction for sex, age, percentage of predicted FEV1, cancer status, batch, and mTIN. The upper and lower hinges correspond to the first and third quartile, the center line represents the median, and whiskers extend from the hinge to the largest or smallest value at most 1.5 times the interquartile range. (**b**) Associations between the expression of ACE2 (tan), CTSL (brown), and TMPRSS2 (olive) and clinical covariates in the nasal and bronchial epithelium. Nasal (pink) and bronchial (purple) brushings were collected from subjects at high-risk of developing lung cancer from 4 different studies (DECAMP bronchus: N = 341; AEGIS bronchus: N = 305; BCLHS bronchus: N = 238; PCA bronchus: N = 133; DECAMP nose: N = 253; AEGIS nose: N =150). The size and color of the bubbles represent the significance and magnitude, respectively, of the t-statistics, calculated using linear modeling of VE gene expression as a function of each clinical variable: smoking status, sex, age, and percentage of predicted FEV1 (and, for the AEGIS dataset, lung cancer diagnosis), correcting for batch and mean Transcript Integrity Number (mTIN). Significance levels: * < 0.05, ** < 0.01, *** < 0.001 (**c**) Expression of VE genes is not significantly correlated (p>0.05, Pearson correlation) between paired AEGIS nasal (x-axis) and bronchial epithelial samples (y-axis) (N = 113). Residual expression values adjusted for sex, age, percentage of predicted FEV1, cancer status, batch, and mTIN were used for the comparison. The blue line is the line of best fit and the grey shading represents the 95% confidence level interval for predictions from the linear model.

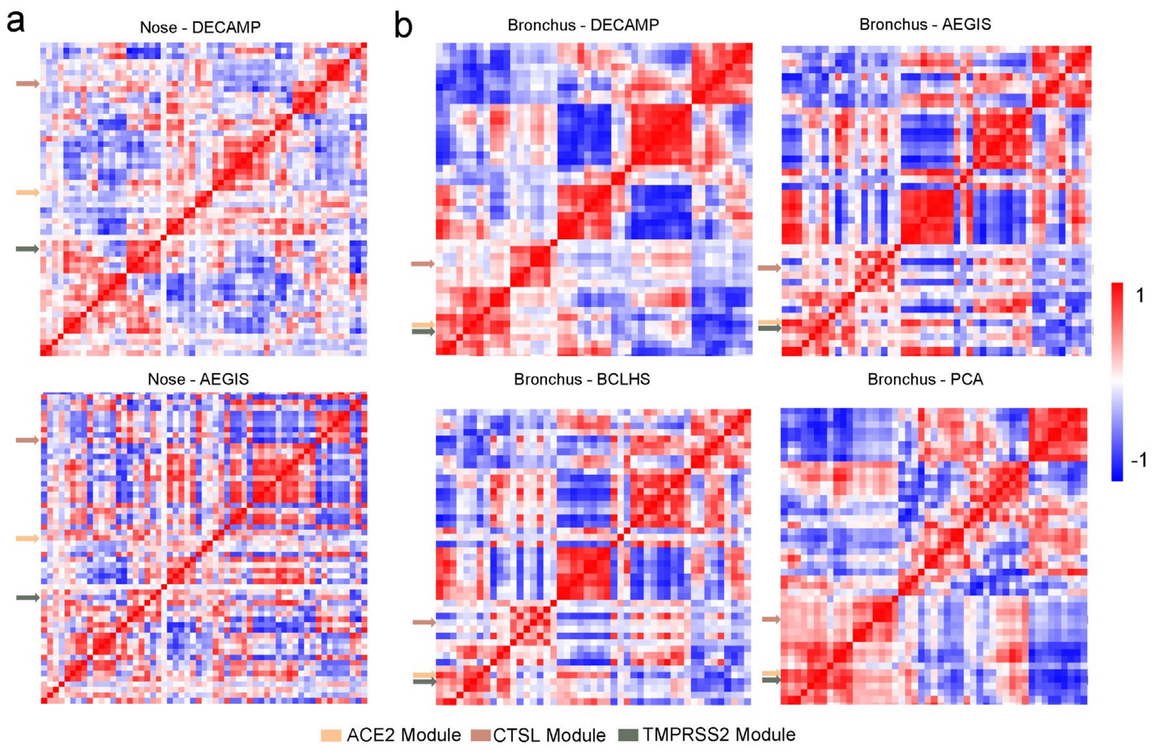


**Figure S2. Modules of co-expressed genes within the nasal and bronchial epithelium.**

Hierarchal clustering of Pearson correlation coefficients between consensus module eigengenes in the (**a**) nasal and (**b**) bronchial epithelium across the cohorts. The modules containing the VE genes are highlighted as follows: ACE2 (tan), CTSL (brown), and TMPRSS2 (olive). The ACE2 and TMPRSS2 modules cluster with one another in the bronchial dataset but not in the nasal dataset. The CTLS module did not cluster with the ACE2 or TMPRSS2 modules in either dataset.

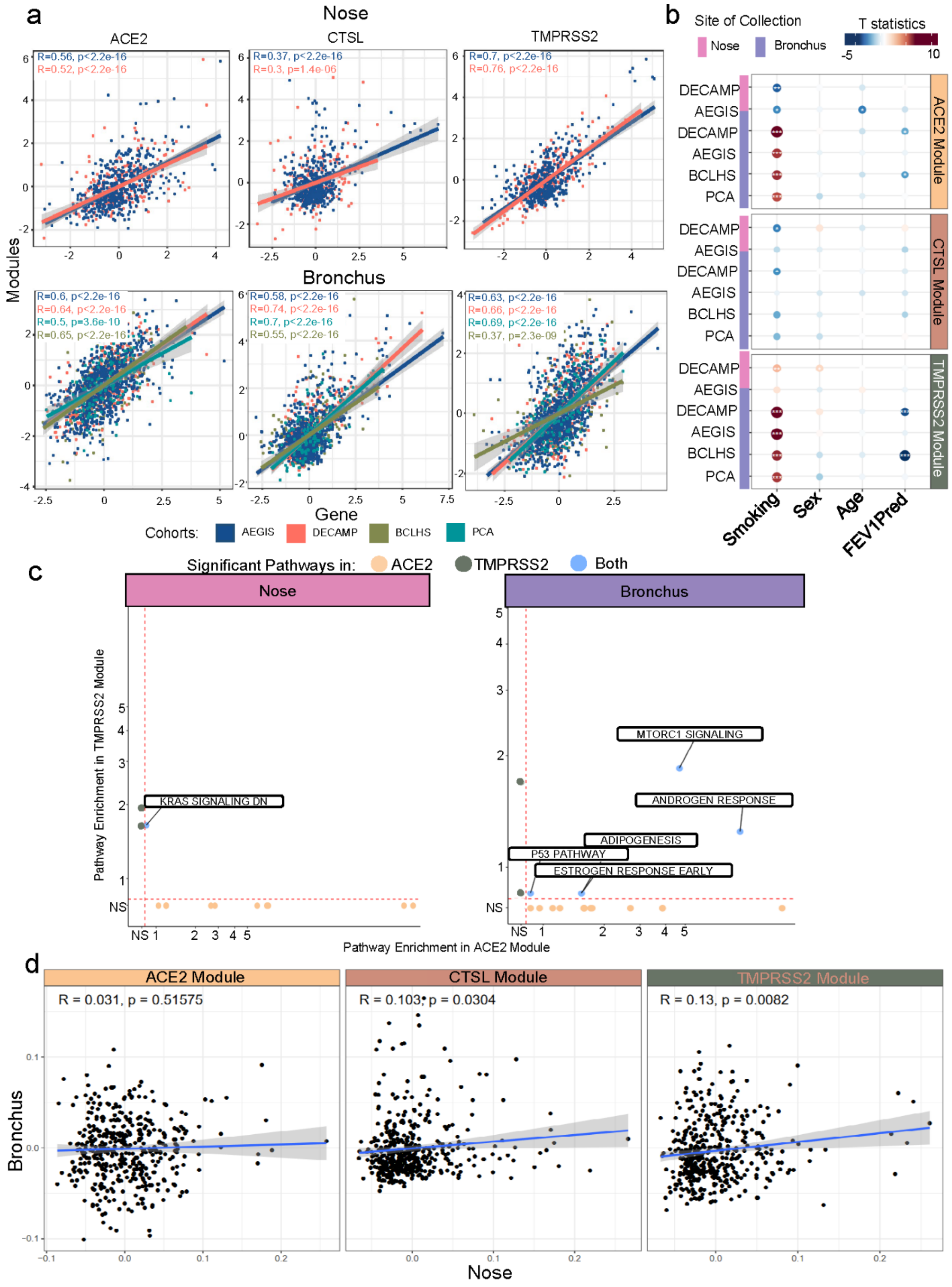
a
Nose
ACE2
CTSL
TMPRSS2
R=0.56, p<2.2e-16
R=0.52, p<2.2e-16
R=0.37, p<2.2e-16
R=0.3, p=1.4e-06
R=0.7, p<2.2e-16
R=0.76, p<2.2e-16
Bronchus
R=0.6, p<2.2e-16
R=0.64, p<2.2e-16
R=0.5, p=3.6e-10
R=0.65, p<2.2e-16
R=0.58, p<2.2e-16
R=0.74, p<2.2e-16
R=0.7, p<2.2e-16
R=0.55, p<2.2e-16
R=0.63, p<2.2e-16
R=0.66, p<2.2e-16
R=0.69, p<2.2e-16
R=0.37, p=2.3e-09
Modules
Gene
Cohorts: AEGIS DECAMP BCLHS PCA
b
Site of Collection
Nose
Bronchus
T statistics
-5
10
DECAMP
AEGIS
DECAMP
AEGIS
BCLHS
PCA
ACE2 Module
CTSL Module
TMPRSS2 Module
Smoking
Sex
Age
FEV1Pred
c
Significant Pathways in: ACE2 TMPRSS2 Both
Nose
Bronchus
Pathway Enrichment in TMPRSS2 Module
KRAS SIGNALING DN
MTORC1 SIGNALING
ANDROGEN RESPONSE
ADIPOGENESIS
P53 PATHWAY
ESTROGEN RESPONSE EARLY
NS
Pathway Enrichment in ACE2 Module
d
ACE2 Module
CTSL Module
TMPRSS2 Module
R = 0.031, p = 0.51575
R = 0.103, p = 0.0304
R = 0.13, p = 0.0082
Bronchus
Nose

**Figure S3. Comparison between the expression of the VE gene modules and their corresponding genes.**

(**a**) The expression of the VE genes is significantly correlated with that of their corresponding gene modules ($p<0.05$, Pearson correlation). Residual expression adjusted for sex, age, percentage of predicted FEV1, batch, and mTIN were used for the comparison. (**b**) Correlations between the expression of VE gene modules and smoking status, sex, age, and percentage of predicted FEV1 across four cohorts with samples collected from nasal (DECAMP N = 211, AEGIS N = 150) and bronchial (DECAMP N = 341, AEGIS N = 305, BCLHS N = 238, PCA N = 133) epithelium. The size and color of the bubbles represent the magnitude and direction, respectively, of the t-statistics calculated using linear modeling of VE gene expression as a function of the clinical variables, correcting for batch and mTIN. Significance level: * < 0.05, ** < 0.001, *** < 0.0001. (**c**) Scatterplots comparing the overrepresentation of MSigDB Hallmark pathway gene sets within the ACE2 and TMPRSS2 gene modules in the nose and bronchus. (**d**) Pearson correlation of the expression of gene modules between paired AEGIS nasal (x-axis) and bronchial (y-axis) epithelial samples (N = 113). Residual expression adjusted for sex, age, percentage of predicted FEV1, cancer status, batch, and mTIN were used for the comparison. The blue line is the line of best fit and the grey shading represents the 95% confidence level interval for predictions from the linear model.

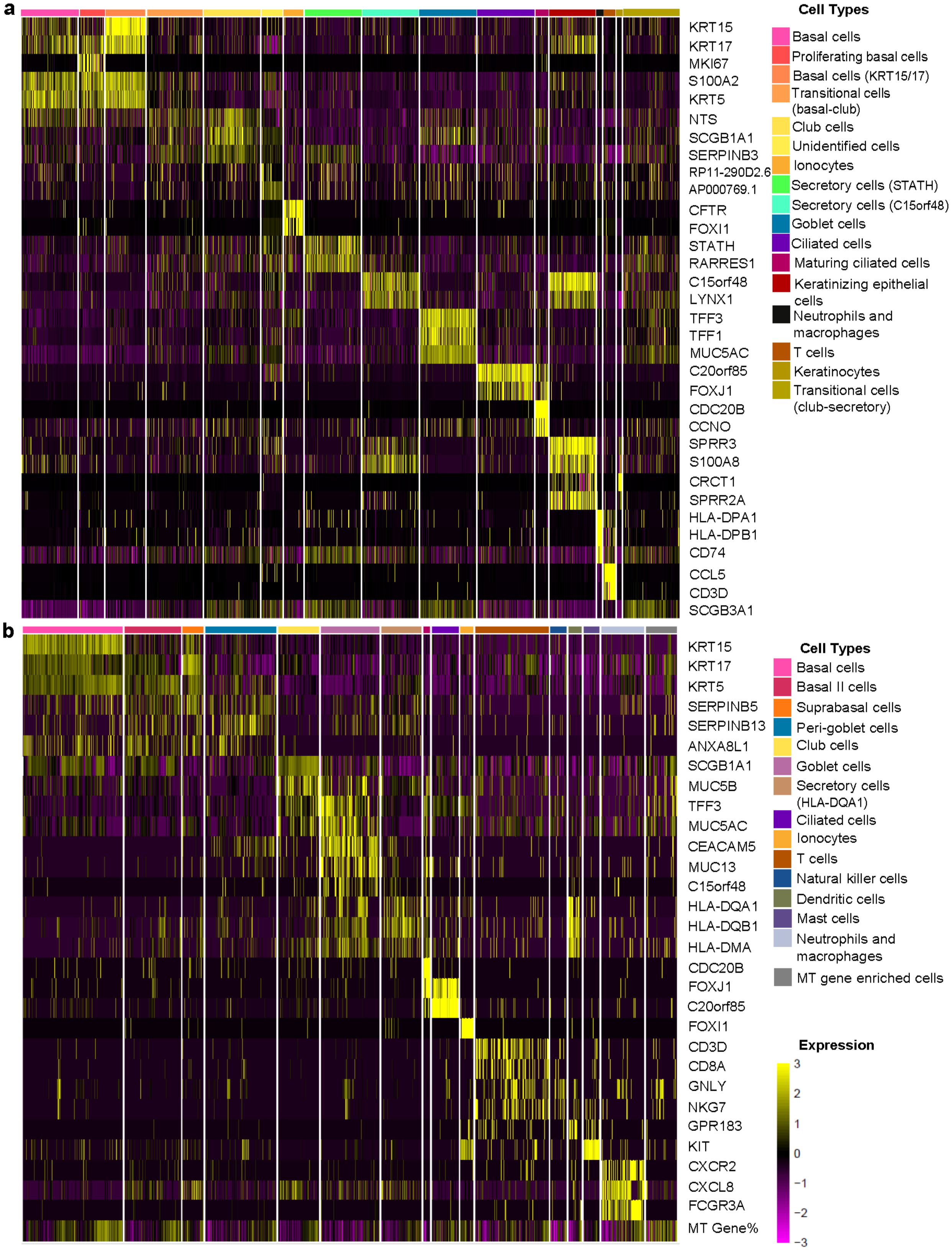
a
KRT15
KRT17
MKI67
S100A2
KRT5
NTS
SCGB1A1
SERPINB3
RP11-290D2.6
AP000769.1
CFTR
FOXI1
STATH
RARRES1
C15orf48
LYNX1
TFF3
TFF1
MUC5AC
C20orf85
FOXJ1
CDC20B
CCNO
SPRR3
S100A8
CRCT1
SPRR2A
HLA-DPA1
HLA-DPB1
CD74
CCL5
CD3D
SCGB3A1
Cell Types
Basal cells
Proliferating basal cells
Basal cells (KRT15/17)
Transitional cells (basal-club)
Club cells
Unidentified cells
Ionocytes
Secretory cells (STATH)
Secretory cells (C15orf48)
Goblet cells
Ciliated cells
Maturing ciliated cells
Keratinizing epithelial cells
Neutrophils and macrophages
T cells
Keratinocytes
Transitional cells (club-secretory)
b
KRT15
KRT17
KRT5
SERPINB5
SERPINB13
ANXA8L1
SCGB1A1
MUC5B
TFF3
MUC5AC
CEACAM5
MUC13
C15orf48
HLA-DQA1
HLA-DQB1
HLA-DMA
CDC20B
FOXJ1
C20orf85
FOXI1
CD3D
CD8A
GNLY
NKG7
GPR183
KIT
CXCR2
CXCL8
FCGR3A
MT Gene%
Cell Types
Basal cells
Basal II cells
Suprabasal cells
Peri-goblet cells
Club cells
Goblet cells
Secretory cells (HLA-DQA1)
Ciliated cells
Ionocytes
T cells
Natural killer cells
Dendritic cells
Mast cells
Neutrophils and macrophages
MT gene enriched cells
Expression
3
2
1
0
−1
−2
−3

**Figure S4. Heatmaps of the expression of the top marker genes within each cell type of the nasal and bronchial epithelium.**

The heatmap displays gene expression patterns for marker genes of each cell population cluster from the **(a)** nasal and **(b)** bronchial epithelium. Markers for each population were found using differential expression testing with the FindMarkers function of the Seurat package (Version 3.0). The expression of each marker gene was z-score normalized across all cells. Individual genes are represented in rows and cells are represented in columns. All bronchial cells and a random subsample of 3,000 nasal cells were used to generate each plot. Yellow indicates high relative gene expression and purple indicates low or no expression. The color bars at the top represent the cell clusters showing in the UMAP of Figure 3.

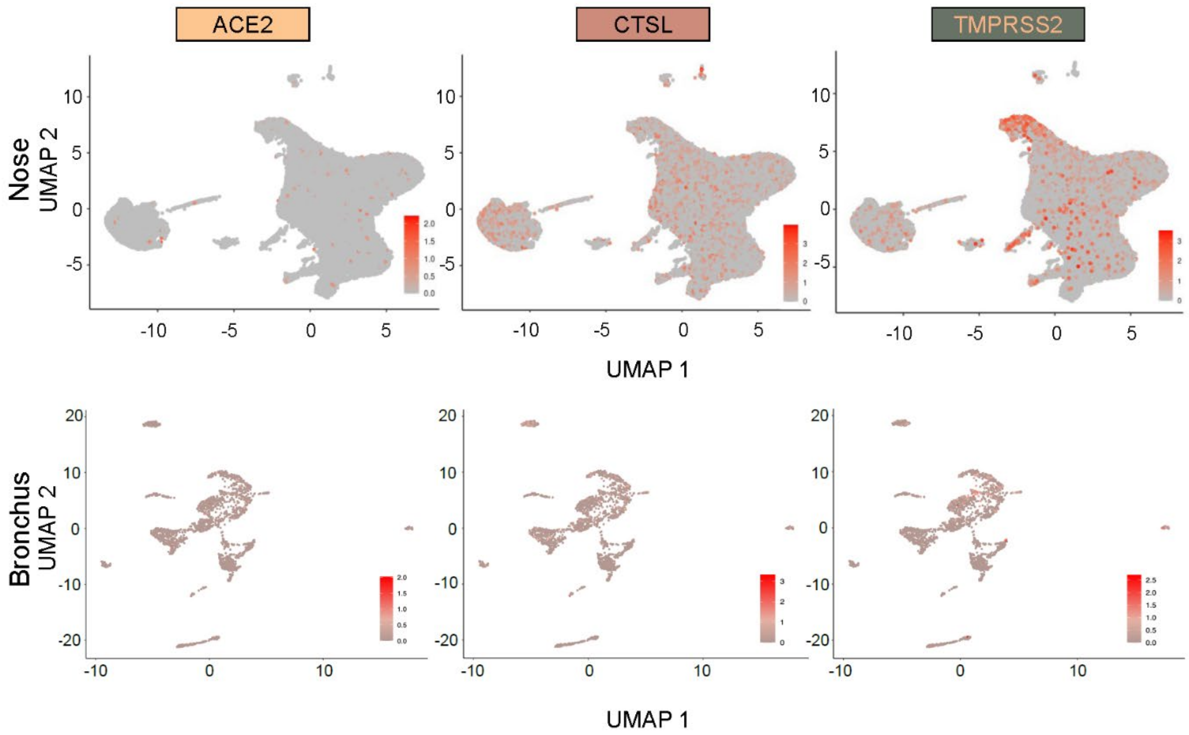


**Figure S5. Sparse expression of individual VE genes in the nasal and bronchial epithelium.**

Expression patterns of the individual VE genes, ACE2 (tan), CTSL (brown), and TMPRSS2 (olive), across different cell types in nasal and bronchial brushing single cell RNA-seq datasets. The same UMAP project shown in Figure 3 was used and the normalized counts of each gene were used for the plot.

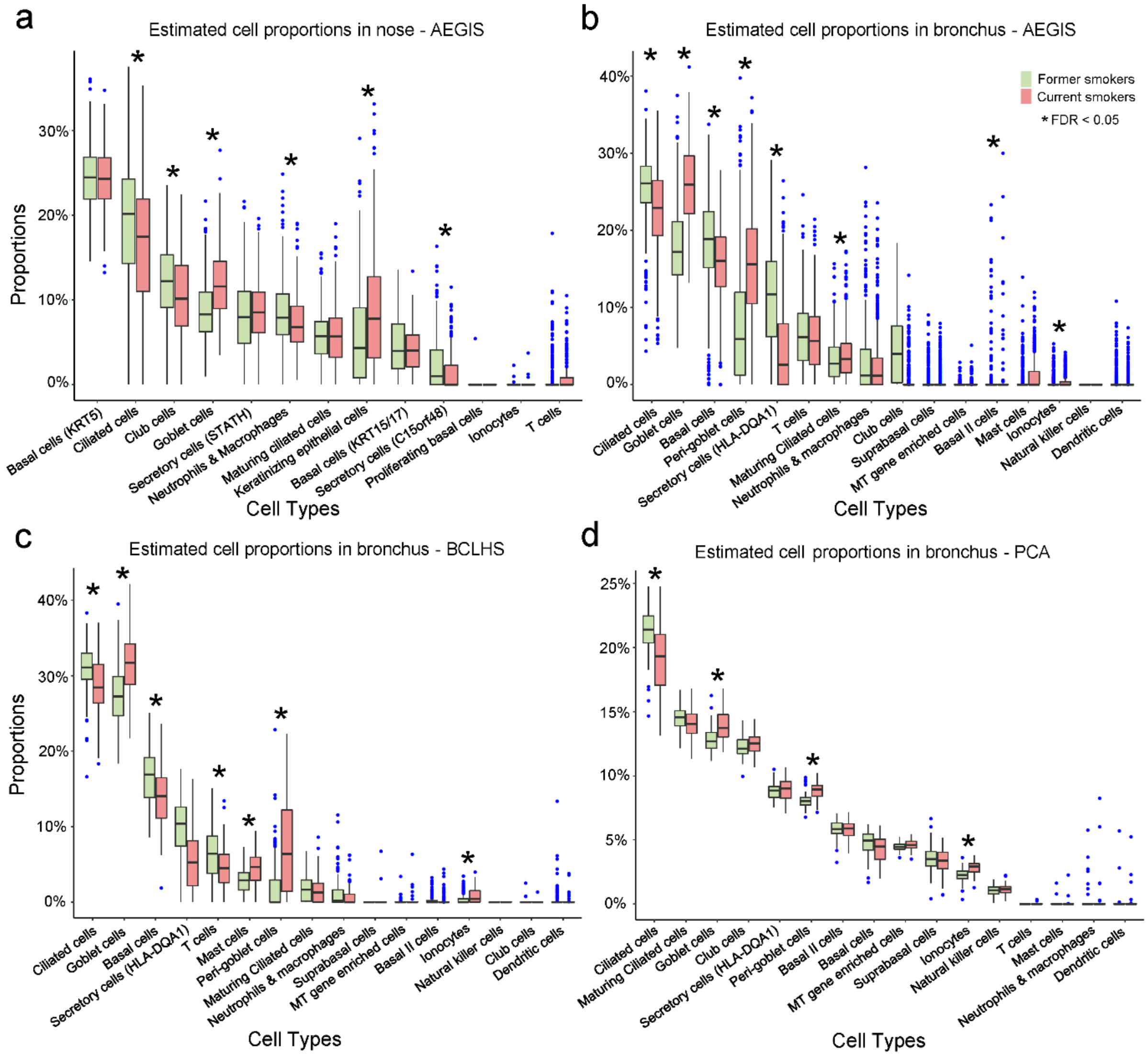


**Figure S6. Cigarette smoking was negatively correlated with the proportion of ciliated cells in both nasal and bronchial epithelium but positively correlated with nasal and bronchial goblet cells, as well as bronchial ionocytes and peri-goblet cells.**

Boxplots of cell proportions estimated by AutoGeneS in the bulk RNA-seq AEGIS (**a**) nasal (N = 508) data, (**b**) AEGIS bronchial (N = 938) data, (**c**) BCLHS bronchial (N = 238) data, and (**d**) PCA bronchial (N = 137) data in current and former smokers. Significant cell proportion differences between current and former smokers were determined by Student's *t* test (* indicates FDR<0.05). The proportion of ciliated cells was significantly lower in current smokers in both nasal and bronchial brushing datasets. The proportion of goblet cells was significantly higher in current smokers in both datasets. In the bronchial brushing dataset, the proportions of ionocytes and peri-goblet cells were also significantly higher in current smokers.

### Supplemental Methods

#### Detailed Description of each Nasal and Bronchial Datasets.

Samples of the DECAMP cohort[1] were collected from ever smokers with indeterminate pulmonary nodules (7-30 mm) in the DECAMP1 study (NCT01785342) and from patients undergoing lung cancer screening in the DECAMP2 study (NCT02504697) (360 bronchial brushings, 288 nasal brushings, and 124 paired samples).

Samples from the AEGIS cohort[2] were collected from patients undergoing bronchoscopy for suspicion of lung cancer and profiled using Affymetrix Human Gene 1.0 ST microarrays (938 bronchial brushings, and 508 nasal brushings, 440 of which are paired samples). The nasal microarray data had previously been deposited in Gene Expression Omnibus (GEO) Series GSE80796.

Samples from the BCLHS cohort (GEO Series GSE37147, n = 238)[3] were collected from bronchial airway brushings from high-risk subjects being followed longitudinally for the development of lung cancer and profiled using Affymetrix Human Gene 1.0 ST microarrays.

Samples from the PCA cohort (GEO Series GSE109743, n = 137)[4] were collected from normal-appearing mainstem bronchus of participants undergoing lung cancer screening by CT scan and profiled using Illumina RNA sequencing. Residual count matrices adjusted for RNA quality (median TIN, mTIN) and batch (Illumina flow cell) were calculated using the original method in the publication and used as input for clinical covariate association and WGCNA analysis.

#### Library Preparation, Sequencing, and Data Preprocessing

DECAMP: Total RNA was isolated from nasal and bronchial brushings using the miRNeasy Mini Kit [Qiagen, Valencia, CA]. RNA integrity was assessed by Agilent BioAnalyzer, and RNA purity was confirmed using a NanoDrop spectrometer. Libraries were generated using the Illumina TruSeq Stranded Total RNA kit and sequenced on Illumina NextSeq 500 and Illumina HiSeq2500 instruments with 75 base-pair paired-end reads (Illumina, San Diego, CA). Demultiplexing and creation of FASTQ files were performed using BaseSpace. We developed an automatic pipeline (https://github.com/compbiomed/RNA_Seq) with standard setups based on the Nextflow framework to obtain the expression levels for each gene[5]. Reads were aligned to the Genome Reference Consortium human build 37 (GRCh37) using STAR[6]. Gene and transcript level counts were calculated using RSEM[7] using Ensembl v75 annotation. Quality metrics were calculated by STAR and RSeQC[8].

AEGIS: Human Gene 1.0 ST CEL files were normalized to generate gene-level expression values using the R environment for statistical computing (version 3.6.0) and the Robust Multiarray Average (RMA) algorithm[9] with an Ensembl Gene-specific probe set mapping (version 22.0.0) from the Molecular and Behavioral Neuroscience Institute (Brainarray) at the University of Michigan [http://brainarray.mbni.med.umich.edu/]. Standardized RMA quality metrics were assessed, including the normalized un-scaled standard (NUSE) error and relative log expression (RLE).

The gene expression profiles were processed before the analysis to adjust for technical effects. Specifically, for DECAMP-nasal and DECAMP-bronchial datasets, normalized counts were calculated by performing variance stabilizing transformation on log2 counts per million (CPM) using DESeq2[10] and batch correction using ComBat-Seq[11]. For the BCLHS dataset, ComBat[12] was used for batch correction.

All samples included for the analysis had sex annotation correlated with the expression of the constitutively expressed Y-linked genes CYorf15A, DDX3Y, KDM5D, RPS4Y1, USP9Y, and UTY. No additional filtering of samples was carried out for calculating clinical correlations. RNA-seq samples with mTIN $\leq$ 60 were not included for generating consensus gene modules.

**Derivation of Consensus Gene Modules and Pathway Enrichment Analysis**

Consensus gene modules were separately identified using the blockwiseConsensusModules function from the WGCNA (Weighted Gene Correlation Network Analysis) package[13] across each dataset. The signed network type was used for both nasal and bronchial derivations. The parameters set for the nasal derivation were a soft power threshold of 12 and a merge cut height of 0.2, and minimum module size of 10. For the bronchial derivation, the parameters set were a soft power threshold of 13, a merge cut height of 0.1, and minimum module size of 30. The Hallmark pathway gene sets from the MSigDB database[14] were leveraged for pathways enrichment analysis (Version v7.2). The implementation of Fisher's exact test within the GeneOverlap package[15] was used to generate p-values and odds ratios to quantify overlap between the MSigDB gene sets and genes in each of the VE modules.

**Single-cell isolation, library preparation, and sequencing**

In the nasal scRNA-seq data, cells were initially dissociated from two nasal swabs [CytoSoft, Camarillo, CA] obtained from the inferior turbinate. The cells were then washed with Phosphate-buffered saline (PBS, Sigma Aldrich, Burlington, MA) and dissociated to single-cell suspensions using 0.25% Trypsin/EDTA (Thermo Fisher, Waltham, MA). Red blood cells were removed after treatment with 1X RBC Lysis Buffer [Thermo Fisher, Waltham, MA] for 2 minutes. Trypan blue exclusion [STEMCELL, Vancouver, BC, Canada] was used for measuring cell viability. The final concentration of cells was measured using a hemocytometer under a light microscope before library preparation using the 10X Genomics Platform [Pleasanton, CA].

In the bronchial scRNA-seq data, the tissue obtained from bronchial brushings was treated with 0.25% Trypsin/EDTA for epithelial sheet dissociation and cells were sorted using a BD FACSAria II. FACS was used to isolate singlet events based on forward scatter height vs. forward scatter area (FSH-H vs. FSH-A). Dead cells (PI+) and red blood cells (GYPA/CD235a+) were stained and excluded. For each donor, single live cells (Hoechst 33342$^{+}$ Propidium Iodide$^{-}$ CD235a$^{-}$) were sorted into two 96-well PCR plates per patient sample.

Both the nasal and bronchial samples were sequenced on an Illumina NextSeq 500 with 75 base-pair single-end reads. Reads were demultiplexed using Cell Ranger (v3.1.0) and aligned to human genome build hg38 (v1.2.0) and tabulated according to unique combination of a Universal Molecular Identifier (UMI) and alignment position.

**scRNA-seq quality control**

Quality metrics and gene-level counts were generated for the single-cell RNA-sequencing samples using Scruff[16]. In the nasal scRNA-seq data, cells had a median of 3095 UMIs, a median of 1277 genes detected, median predicted contamination of 0.04%, and 7.5% predicted doublets. Cells were identified as poor quality if they satisfied any of the following criteria: 1) bottom quantile for a total number of genes detected, 2) bottom quantile for total library size and 3) greater than

30% the counts mapped to the mitochondrial genome. In total, 15787 cells were filtered and 34833 cells were kept for further analysis.

In the bronchial scRNA-seq data, cells had a median of 4580 UMIs, a median of 1870 genes detected, median predicted contamination of 0.168 %, and 7.51% predicted doublets. Cells were identified as poor quality if they met any of the following criteria: 1) bottom quantile for a total number of genes detected, 2) bottom quantile for total library size and 3) top quantile for the percentage of counts mapped to the mitochondrial genome. Overall, 1189 cells (including 34 empty wells) were filtered and 2075 cells were kept for further analysis.

**Cell Type Identification**
Cells were defined by examining the relative expression level of knowledge-based gene markers. Transitional cells were named based on their relative location on the UMAP plot and expression of a combination of marker genes.

In the nasal scRNA-seq data, markers included KRT5, KRT15 and KRT 17 (basal cells), SCGB1A1 (club cells), MUC5AC (goblet cells), FOXJ1 (ciliated cells), FOXI1 (ionocytes), CD3D (T cells), CD74 (neutrophils and macrophages), STATH and C15orf48 (secretory cells), CDC20B (maturing ciliated cells), and SPRR3 (keratinizing epithelial cells). In the bronchial scRNA-seq data, markers included KRT5 (basal cells), CEACAM5 (peri-goblet cells), SCGB1A1 (club cells), MUC5AC (goblet cells), FOXJ1 (ciliated cells), FOXI1 (ionocytes), CD3D (T cells), GNLY (NK cells), KIT (mast cells), and CXCR2 (neutrophil and macrophages).

**Deconvolution of bulk RNA-seq gene expression using cell type markers derived from the scRNA-seq datasets.**
To dissect cell population proportions from bulk RNA samples, reference gene expression profiles (GEPs) were derived using the nasal and bronchial scRNA-seq data following a similar procedure. Marker genes of each cell type were identified using the FindMarkers function within the Seurat package using the MAST method of modeling (FDR < 0.05, logFC > 0.25). The GEPs of each cell population were defined as the mean expressions of the corresponding cell cluster using Seurat scaled counts. After generating the GEPs of the cell populations, the AutoGenes function within the Autogenes package[17] was called to further identify the top 400 most informative genes for deconvolution using multi-objective optimization. The filtered informative genes GEPs were defined as the signature matrix. Nu Support Vector regression was employed to deconvolute or predict cell proportions from the bulk RNA-seq data based on the filtered informative genes. For the DECAMP and PCA datasets, normalized and batch corrected counts were used, and for the BCLHS dataset, batch corrected array measurements were used for the deconvolution.
